# Attitude and Perceived Barriers towards Research among Undergraduate Medical Students of Bangladesh

**DOI:** 10.1101/2021.04.30.21256373

**Authors:** Jannatul Ferdoush, Fatema Johora, IkramUllah Khan, Sharif Mohammad Towfiq Hossain, Halima Sadia, Fatiha Tasmin Jeenia, Sameera Shafique Chowdhury, Nagina Sultana, Shagorika Sharmeen

## Abstract

**Background:** Undergraduate research opportunities teach students not only how to conduct research, but they too learn problem-solving aptitudes. Participating in research also increases students’ interest is being involved and making special contributions to the academic field. Therefore, the aim of our study was intended to assess the attitude and perceived barriers toward research among the medical undergraduates of Bangladesh.

**Methods:** A cross-sectional questionnaire survey was conducted among third, fourth, and fifth year students across medical colleges in Bangladesh, during the month of July, 2020 to December, 2020. A Google-linked questionnaire was disseminated to the students via different social platform and the responses were received through Google drive.

**Result:** The questionnaire survey received responses from 1279 students, with 94% claiming to be familiar with the scientific method. 82.7% of students mentioned they could design and execute a research project as well as can write scientific articles. More than half of the respondents (66.4%) expressed an interest in participating in research. Almost all respondents (96.7%) agreed that research is beneficial as it aid critical thinking and policy implementation. 79.8% of respondents opined that education on research methodology should be required in the medical curriculum. Majority of the respondents reported that lack of time and priorities to do research work because of compact academic tasks (89.1%), insufficient guidance (86.6%), lack of familiarities with research methodology (87.5%) and statistical analysis (85.2%) are the barriers of research.

**Conclusion:** Our findings indicated that Bangladeshi medical students have a positive attitude toward research and that research methodology should be taught in undergraduate medical education. In order to increase participation in research, the challenges identified by students should be addressed.

## Introduction

There is a constant influx of new knowledge in today’s world where new ideas are emerging, and old ideas are being reshaped or replaced. Physician need to learn how to verify authentic information in this era of information epidemics. In order to advance the medical profession, research is essential in this age of evidence-based medicine to offer the best possible treatment to the patient [1]. There is a growing concern about research in both developing and developed countries, as medical research has the potential to improve healthcare [2]. Access to research experience at undergraduate level could be crucial for future physicians as they will be acquainted with the process of generating scientific evidence as well as the challenges and health issues that people, societies, and health systems face requires scientific study from the early days of their career, and it might be reflected throughout their life. Students engaged in research develop specific research skills, self-confidence, self-directed learning, critical thinking, and communication skills [3, 4]. On the other hand, incorporation and inclusion of medical students in basic research could be a way to address the shortage of medical scientists. [5]

Over the last three decades, efforts to encourage undergraduate research have increased. In 1978, the United States established the Council on Undergraduate Research (CUR) [6]. According to “outcomes for Graduates 2018,” the General Medical Council (GMC) found research to be a top priority in medical education [7]. Student-selected components (SSCs) that are used in the curriculum of many UK medical schools with the aim of increasing student exposure to research [8]. The Medical Research Volunteer Program (MRVP) was established in Lebanon 2014 to assist undergraduate students participating in medical research [9]. Here in Bangladesh, research activities are still in its infancy [10]. While research methodology are included in undergraduate programs in many countries,[11] but not included in medical curricula of Bangladesh and many other developing countries [12]. Bangladesh International Medical Students’ Science Congress (BIMSSCON) has promoted scientific education among medical students of Bangladesh since its inception in 2015, by providing a forum for them to exchange ideas, information, and improves networking [13]. IFMSA Bangladesh (The International Federation of Medical Students Associations) is a medical student-run organization in Bangladesh. Since 2015, it has been involved in activities such as SCOREsearch, a three-month training program on the fundamentals of research methodology designed to give students a hands-on approach and help them to develop their capacity [14].

Though students have a positive attitude toward science, they need a supportive and positive atmosphere in order to enhance their research knowledge, develop skills, and resolve the barriers of conducting scientific research [15]. Several studies have shown that the presence of research obstacles contributes to differences in research theory and conduct [16]. Lack of proper knowledge and skills in scientific method and statistics and lack of interest in research, lack of time as set extra burden, lack of training, lack of reward, lack of motivation, and lack of supervision all are identified as obstacles in scientific research [17, 18]. In most cases, students are reluctant to participate in research due to lack of scientific training programs and inadequate supervision [18]. A positive research experience requires the role of a supervisor and a good relationship with the supervisor [19]. The huge curriculum and lack of time in medical education is the main reason why students are less involved in research. As a result, they do not have the skills to write protocols in post graduation.

It is possible to improve students’ awareness and attitude toward research by making research projects mandatory in undergraduate curriculum as part of research methodology [18, 20]. It is important to understand undergraduate students’ attitudes towards research and the obstacles they face in conducting research. Current study was conducted with the aim to assess the attitude and perceived barriers toward research among the medical undergraduates of Bangladesh in order to intervene and take the appropriate measures to overcome the obstacles by recognizing them in upcoming days.

## Materials and Methods

### Study design and population

The cross sectional questionnaire survey was conducted among the 3^rd^, 4^th^ and 5^th^ year medical student across the medical colleges of Bangladesh during the month of July, 2020 to December 2020. Ethical approval was taken from institutional review board (IRB) of BGC trust medical college, Chattogram. The student was informed about their participation is voluntary and to maintain the confidentiality the response was anonymous.

### Study procedure

A Google-linked questionnaire was developed based on literature search and pilot test were carried out to 30 medical students of 3^rd^, 4^th^ and 5^th^ year to check its clarity and reliability. However, the responses were not included in this survey.

The questionnaire was disseminated to the student via different facebook Page and Groups of medical students of Bangladesh, whats app, messenger, and other social platform. Few students volunteer were selected from different medical colleges to distribute the questionnaire and properly address the study. The responses were received through Google drive. The automated Google form received only one response from one email address to ovoid duplicate or multiple responses from a single respondent.

### Outcome Measure

1. To identify undergraduate medical students’ attitude towards research such as interest to participate in research work, familiarities with scientific methods, value of medical research and self-rated competency on study design and write scientific paper.
2. Students perceived barrier towards research such as lack of time, priorities and interest to do research, lack of familiarities with research methodologies and statistical analysis and insufficient guidance.

## Result

A total of 1279 student were responded to the questionnaire survey. Among them, 63.3% were female and 36.7% were male students. Response rate by academic year were 37%, 36.3%, and 26.7% from 3^rd^, 4^th^ and 5^th^ year students respectively.

**Fig 1:**
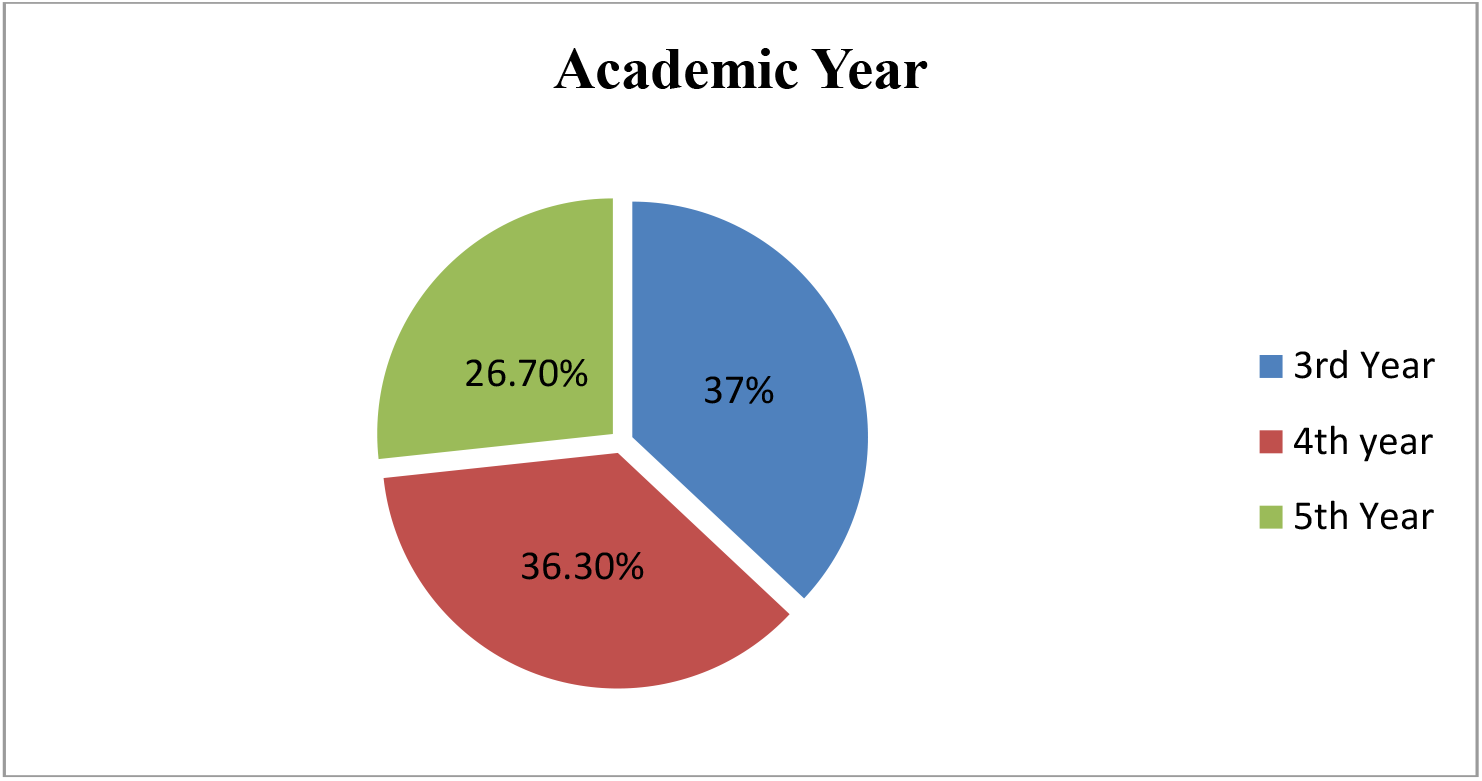
Distribution of undergraduate medical student according to academic year.

Table I showed that 94% students responded that they are familiar with scientific method. 82.7% students mentioned that they can design and perform a research project and write scientific paper. More than half of the respondents (66.4%) interested to participate in research. Almost all respondent (96.7%) agreed that research is beneficial as it is helpful for critical thinking and policy implementation. 79.8% respondent opined that education on research methodology should be compulsory in the medical curriculum.

**Table 1:** Undergraduate medical students’ attitude towards research.

| Attitude towards Research | Response(n=1279)<br>F=63.3% M= 36.7% |  |  |
| --- | --- | --- | --- |
|  | Totally agree/agree | Disagree | No comments |
| Familiarity with the scientific method | 94% | 2.1% | 3.8% |
| Design & perform a research project and write scientific paper | 82.7% | 7.8% | 9.6% |
| Interested to participate in research work | 66.4% | 14.2% | 30.8% |
| Research is beneficial as it is helpful for critical thinking and policy implementation | 96.7% | 0.4% | 2.7% |
| Education on Research Methodology should be compulsory in the medical curriculum | 79.8% | 5.4% | 14.9% |

Majority of the respondents (89.1%) mentioned that lack of time and priorities to do research work because of compact academic tasks, lack of familiarities with research methodology (87.5%) and statistical analysis (85.2%) are the barrier of research. Insufficient guidance and lack of interest to do research also mentioned by 86.6% and 57.4% responded respectively. (Table II).

**Table II:**
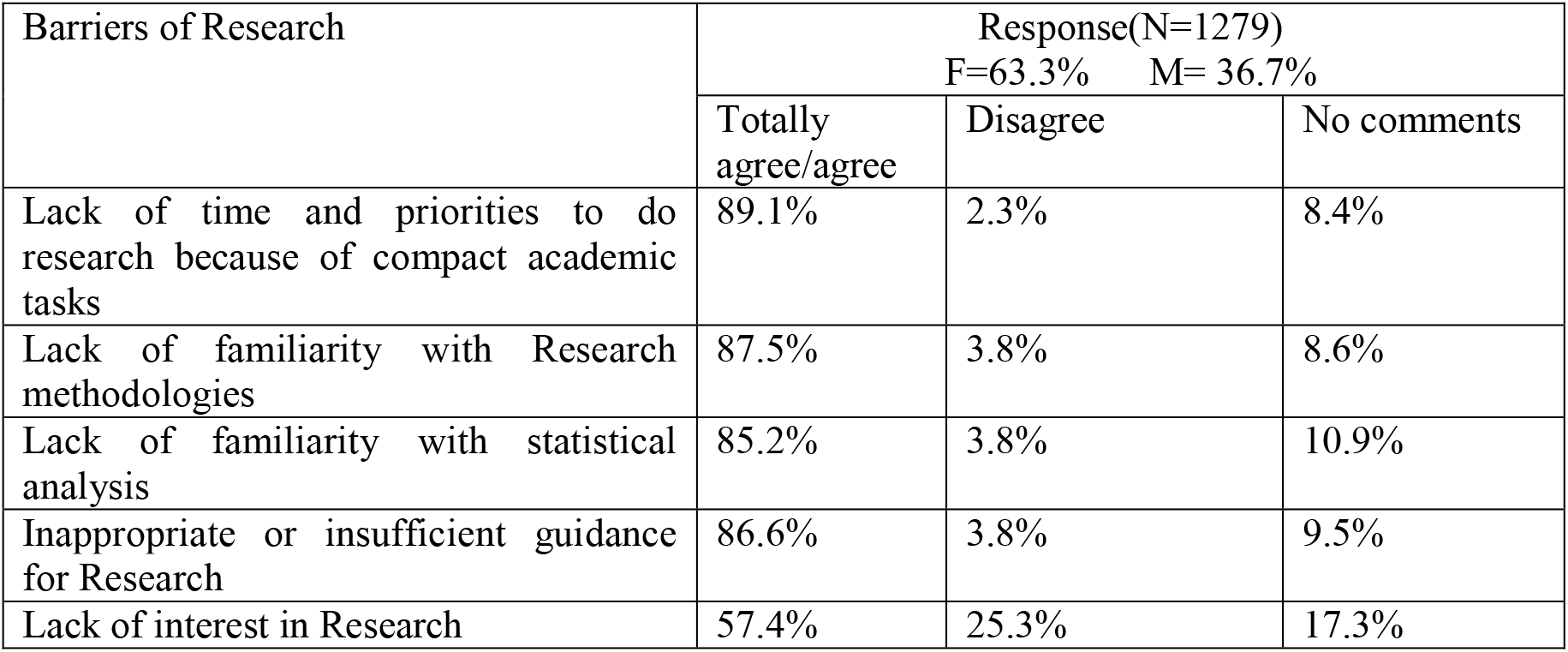
Undergraduate medical students’ perceived barriers of research.

## Discussion

Early introduction to scientific research at the undergraduate level plays a noteworthy role in constructing an advanced medical education for students [21]. Undergraduate Research is the driving force behind a student’s interest in what they are studying and thinking about. This is more than simply memorizing facts and figures for an exam and then forgetting about them until the exam is over. Conducting research necessitates sufficient awareness, a positive mindset, and the identification of research barriers [22]. The aim of our survey was to determine undergraduate medical students’ attitudes toward research and perceived barriers to conducting research.

Medical students with previous scientific research understanding usually have a more positive attitude regarding research [23]. According to our findings, the majority of students (94%) are familiar with the scientific method. This result is satisfactory. Our findings contradict the findings of an earlier study by Osman T et al. (2020) in which half of the students reported a lack of familiarity with research [21]. Almost two-thirds of respondents self-rated their abilities to design and carry out research projects, as well as write scientific papers. In contrast to our findings, the ratter et al study (2018) found that undergraduate medical students rated their competencies as inadequate; especially in statistics and scientific writing [24]. Several studies have shown that students misjudge their competencies and believe they are under-trained [25]. In our survey, more than half of the respondents (66.4%) expressed an interest in participating in research. Several studies have found similar results, with students expressing a positive attitude toward research [24, 26, 27]. Students already have a high degree of stress as a result of the overburdened medical curriculum, and they do not have sufficient room to do research or spend time on research [28]. Therefore it is essential that design and content of the medical curriculum should reallocate time that students have enough time on research.

The most significant characteristics of undergraduate research are critical thinking and problem-solving abilities. Medical students should be encouraged to conduct research and improve their analytic skills, which are in high demand right now [29]. According to our findings, undergraduate students believe that research is beneficial and useful for critical analysis and policy implementation. Similar findings were found in the Houlden et al. (2016) study, where students appreciated the role of research in improving critical thinking when carrying out research projects during their study period [30]. Tomorrows Doctors, the Scottish Deans Medical Education Group, and by the Manual for Good Medical Practice USA all strongly recommend encouraging research-specific skills as well as the development of critical thinking, time management, communication skills, and teamwork among undergraduate medical students. [29, 31-33]. In the current survey, more than two-thirds (79.8%) of students thought that research methods should be taught in the undergraduate curriculum. A study also reported that, 60% of medical students agreed to include research methodology in their medical curriculum [34]. The rationale for including research methods in medical curricula may be to facilitate research experience during medical studies. This will foster a positive attitude toward research and critical appraisal of the medical literature, as well as encourage interest in medical research as an academic career [12].

Our study attempted to investigate the research barriers mentioned by Bangladeshi medical students. The majority of students cited that lack of time and priorities for conducting research as a result of their compact academic workload. The lack of familiarity with scientific methods and statistical analysis is cited as barriers to research by 87.7 % of students. This might be due to insufficient emphasis given in research methodology and statistical analysis during medical study. To assist undergraduate students, medical college can host a seminar on scientific research to educate them and clear up any misconceptions about the nature and value of scientific research [26]. Consistent with our findings Pallamparthy S et al.(2019) study found that students stated lack of knowledge (53%), lack of interest (54 %), financial grants (62%), and lack of time as obstacles to research (59%) [34]. Another study by El Achi D et al. (2020) found that 56% student believe research is not a waste of time and does not interfere with studies, even though some medical students believe their courses are more important than allocating time for medical research [24]. Similar barriers to research were also explored in studies from various countries [15, 16, 18].

In the current study, 86.6% stated that insufficient guidance is an important barrier of research. This might be due to medical faculty or principal investigators pay less emphasis on allocating undergraduate students [26]. According to the findings of the Funston et al.(2019) study, students face a lack of supervision and direction while conducting research [35]. In fact, according to a study conducted by Ahn J et al. (2007) pointed out that undergraduate students are concerned about finding the right balance between clinical and academic education [36]. Appropriate feedback by teacher can facilitate students to have a positive research mindset. Medical educators should place a greater focus on the challenges identified by students, such as mentoring and guidance, in order to improve undergraduate medical students’ research experiences [26].

### Conclusion

Undergraduate medical students of Bangladesh were enthusiastic about conducting medical research. The majority of medical students of Bangladesh believe that research methods should be included in the undergraduate curriculum. Due to condensed academic activities; there is a lack of time and goals for research work, as well as a lack of familiarity with research methods, statistical analysis, and poor guidance. Our findings may be used to improve undergraduate student research in medical education and as a recommendation for overcoming obstacles.

## Data Availability

https://docs.google.com/forms/d/e/1FAIpQLSfZfvZEvVtfp2yGwtlUWBsFs-K12_0xwtqY_As_kXPLvvLiCw/viewform?usp=sf_link

## Notes

### Competing Interest Statement

The authors have declared no competing interest.

### Funding Statement

None

### Author Declarations

Institutional Review Board Of BGC Trust Medical College.

